# Temporal changes in SARS-CoV-2 clearance kinetics and the optimal design of antiviral pharmacodynamic studies: an individual patient data meta-analysis of a randomised, controlled, adaptive platform study (PLATCOV)

**DOI:** 10.1101/2024.01.16.24301342

**Authors:** Phrutsamon Wongnak, William HK Schilling, Podjanee Jittamala, Simon Boyd, Viravarn Luvira, Tanaya Siripoon, Thundon Ngamprasertchai, Elizabeth M Batty, Shivani Singh, Jindarat Kouhathong, Watcharee Pagornrat, Patpannee Khanthagan, Borimas Hanboonkunupakarn, Kittiyod Poovorawan, Mayfong Mayxay, Kesinee Chotivanich, Mallika Imwong, Sasithon Pukrittayakamee, Elizabeth A Ashley, Arjen M Dondorp, Nicholas PJ Day, Mauro M Teixeira, Watcharapong Piyaphanee, Weerapong Phumratanaprapin, Nicholas J White, James A Watson, the PLATCOV Collaborative Group

## Abstract

**Background:** Effective antiviral drugs prevent hospitalisation and death in COVID-19. Antiviral efficacy can be assessed efficiently in-vivo by measuring rates of SARS-CoV-2 clearance estimated from serial viral genome densities quantitated in nasopharyngeal or oropharyngeal swab eluates. We carried out an individual patient data meta-analysis of unblinded arms in the PLATCOV platform trial to characterise changes in viral clearance kinetics and infer optimal design and interpretation of antiviral pharmacometric evaluations. PLATCOV is registered at ClinicalTrials.gov, NCT05041907.

**Methods:** Serial viral density data were analysed from symptomatic, previously healthy, adult patients (within 4 days of symptom onset) enrolled in a large multicentre randomised adaptive pharmacodynamic platform trial (PLATCOV) comparing antiviral interventions for SARS-CoV-2. Viral clearance rates over one week were estimated under a hierarchical Bayesian linear model with B-splines used to characterise temporal changes in enrolment viral densities and clearance rates. Bootstrap re-sampling was used to assess the optimal duration of follow-up for pharmacometric assessment, where optimal is defined as maximising the expected z-score when comparing effective antivirals with no treatment.

**Results:** Between 29 September 2021 and 20 October 2023, 1262 patients were randomised. Unblinded data were available from 800 patients (16,818 oropharyngeal viral qPCR measurements) of whom 63% (504/800) were female. 98% (783/800) had received at least one vaccine dose and over 88% (703/800) were fully vaccinated. SARS-CoV-2 viral clearance was biphasic (bi-exponential). The first phase (*α*) was accelerated by effective interventions. For all the effective interventions studied, maximum discriminative power (maximum expected z-score) was obtained when evaluating serial data from the first 5 days after enrolment. Over the two-year period studied, median viral clearance half-lives estimated over 7 days have shortened from 16.6 hours (interquartile range [IQR]: 15.3 to 18.2) in September 2021 to 9.2 hours (IQR: 8.0 to 10.6) in October 2023 in patients receiving no antiviral drugs, equivalent to a relative reduction of 44% [95% credible interval (CrI): 19 to 64%]. A parallel trend was observed in treated patients. In the 158 patients randomised to ritonavir-boosted nirmatrelvir (3,380 qPCR measurements), the median viral clearance half-life declined from 6.4 hours (IQR: 5.7 to 7.3) in June 2022 to 4.8 hours (IQR: 4.2 to 5.5) in October 2023, a relative reduction of 26% [95%CrI: –4 to 42%].

**Conclusions:** SARS-CoV-2 viral clearance kinetics in symptomatic vaccinated individuals have accelerated substantially over the past two years. Antiviral efficacy in COVID-19 can now be assessed efficiently in-vivo using serial qPCRs from duplicate oropharyngeal swab eluates taken daily for 5 days after drug administration.

**Funding:** Wellcome Trust Grant ref: 223195/Z/21/Z through the COVID-19 Therapeutics Accelerator.

## Background

Effective SARS-CoV-2 antivirals taken early in the course of COVID-19 illness accelerate viral clearance, hasten symptom resolution, reduce transmission, and lower the probability of progression to severe disease [1–4]. Several small molecule drugs and monoclonal antibodies have proven antiviral efficacy in COVID-19, although monoclonal antibodies are no longer used widely as immune evasion resulting from viral evolution has reduced or abrogated their antiviral effects. Currently the most effective approved small molecule antiviral drug is ritonavir-boosted nirmatrelvir, a main (3C-like) protease inhibitor [5]. Nirmatrelvir reduces progression to severe disease in an unvaccinated high-risk population by around 90% [3]. But the combination drug is expensive, ritonavir is contraindicated in many individuals because of drug-drug interactions, and ritonavir-boosted nirmatrelvir frequently results in troubling dysgeusia [3]. The development of better tolerated drugs (for example ensitrelvir, also a main protease inhibitor [6]) which could be administered more widely would be of considerable public health value, particularly if they were affordable. To guide policies and practices the antiviral activities of new drugs need to be compared against current treatments. Antiviral interventions can be assessed and compared using acceleration in the rate of viral clearance as a surrogate for clinical benefit [7–9]

The natural history of SARS-CoV-2 infection has changed markedly over the past four years since the beginning of the pandemic [10]. Serious clinical outcomes, notably life-threatening inflammatory pneumonitis, are now very rare. As a result, it has become very difficult to demonstrate clinical efficacy for new antiviral drugs because the required trial sample sizes have become prohibitively large. This was illustrated in the very large PANORAMIC trial of molnupiravir in the UK where only 203 primary events were observed in >25,000 randomised at-risk patients [11]. An alternative approach is to use rates of in-vivo viral clearance to characterise and compare antiviral efficacies [12]. This is relatively straightforward and requires orders of magnitude fewer patients [13]. PLATCOV is an ongoing multicentre phase 2 adaptive, open label, randomised, pharmacometric platform trial in symptomatic low risk adults with COVID-19 (NCT05041907) [13]. Results from this trial have demonstrated the utility of this approach in identifying ineffective drugs, and assessing and comparing those which are clinically effective [5, 13–16].

Viral clearance in COVID-19 follows an approximate bi-exponential (biphasic) decay pattern [17–19]. Previous studies have shown that effective antiviral interventions increase the rate of viral clearance in the first phase [12, 20]. The effect of antivirals on the second phase is less clear and of lesser importance as viral densities are usually fairly low (i.e., unlikely to be transmissible), close to the limit of detection, and clear spontaneously in individuals who are not immunocompromised. The majority of small molecule drugs are given for up to 5 days (e.g., remdesivir, molnupiravir, ritonavir-boosted nirmatrelvir) and have short elimination half-lives. Thus, the primary aim of the PLATCOV trial was to characterise and compare antiviral effects during the first phase of viral clearance. For this reason, the primary endpoint included measured viral densities only up until day 7. In this paper we present an analysis of viral clearance in all patients with unblinded data in the PLATCOV study in order to characterise temporal changes in viral kinetics and re-assess the optimal approach for characterising and comparing antiviral effects in-vivo.

## Methods

### The PLATCOV trial

PLATCOV is an ongoing, open-label, multicentre, phase 2, randomised, controlled, adaptive pharmacometric platform trial in Thailand, Brazil, Pakistan, and Laos. The trial provides a standardised quantitative comparative method for in-vivo assessment of potential antiviral treatments in adults at low risk with early symptomatic COVID-19. The primary endpoint is the rate of viral genome clearance estimated under a linear model fitted to the log viral load (measured by qPCR in daily duplicate oropharyngeal viral swab eluates) data currently sampled on Day 0 and over 7 days of follow-up (8 days in total), denoted *α*_0−7_. All patients receive symptomatic treatment (mainly paracetamol).

PLATCOV is coordinated and monitored by the Mahidol Oxford Tropical Medicine Research Unit (MORU) in Bangkok. The trial was overseen by a trial steering committee and was conducted according to Good Clinical Practice principles. PLATCOV is registered at ClinicalTrials.gov, NCT05041907.

#### Ethics statement

The trial was approved by the Oxford University Tropical Research Ethics Committee (Oxford, UK) and ethics committees in each country. The results were reviewed regularly by a data and safety monitoring board. In Thailand the trial was approved by the Faculty of Tropical Medicine Ethics Committee, Mahidol University, (reference TMEC 21-058); in Brazil by the Research Ethics Committee of the Universidade Federal de Minas Gerais (COEP-UFMG, Minas Gerais, Brazil, COEP-UFMG) and National Research Ethics Commission-(CONEP, Brazil, COEP-UFMG and CONEP Ref: CAAE:51593421.1.0000.5149); in Laos by the National Ethics Committee for Health Research (NECHR, Lao People’s Democratic Republic, Submission ID 2022.48) and the Food & Drugs Department (FDD, Lao People’s Democratic Republic, 13066/FDD_12Dec2022); in Pakistan by the National Bioethics Committee (NBC No.4-87/COVID-111/22/842) the Ethics Review Committee (ERC 2022-7496-21924) and the Drug Regulatory Authority (DRAP Ref: No.0318/2022-CT (PS)).

#### Participants

Eligible participants were previously healthy adults aged 18–50 years who gave fully informed consent for full participation in the study. The entry criteria were: (i) SARS-CoV-2 positive as defined either as a nasal lateral flow antigen test that became positive within 2 minutes (STANDARD Q COVID-19 Ag Test, SD Biosensor, Suwon-si, South Korea) or a positive PCR test with a cycle threshold value less than 25 (all viral gene targets) within the previous 24 h (both these tests ensure the majority of recruited patients have high viral densities); (ii) reported symptoms of COVID-19 for less than 4 days (<96 h); (iii) oxygen saturation on room air ≥96% measured by pulse oximetry at the time of screening; (iv) unimpeded in activities of daily living; (v) agreed to adhere to all procedures, including availability and contact information for follow-up visits.

Exclusion criteria included taking any concomitant medications or drugs, chronic illness or condition requiring long-term treatment or other clinically significant comorbidity, laboratory abnormalities at screening (haemoglobin <8 g/dL, platelet count <50 000/µL, abnormal liver function tests, and estimated glomerular filtration rate <70 mL/min per 1·73 m^2^), pregnancy (a urinary pregnancy test was performed in females), actively trying to become pregnant, lactation, contraindication or known hypersensitivity to any of the proposed therapeutics, currently participating in a COVID-19 therapeutic or vaccine trial, or evidence of pneumonia (although imaging was not required). After a detailed explanation of study procedures and requirements all patients provided fully informed written consent.

Block randomisation was performed for each site via a centralised web-based application. At enrolment, after obtaining fully informed consent and entering the patient details, the app provided the study drug allocation. The no study drug group (unblinded, no placebos were used) comprised a minimum proportion of 20% of patients at all times, with uniform randomisation ratios applied across the active treatment groups. The laboratory team were masked to treatment allocation and the clinical investigators were masked to the virology results until the study group was terminated. Apart from the trial statisticians (JAW and PW), the clinical investigators were all masked to the quantitative PCR (qPCR) results.

Patients were included in this analysis if they had been randomised to a currently unblinded treatment arm and had at least two days of follow-up (i.e., sufficient to estimate a clearance slope).

#### Procedures

All treatments were directly observed. Oropharyngeal swabs were taken as follows by trained study nurses. A flocked swab (Thermo Fisher MicroTest® [Thermo Fisher, Waltham, MA, USA] and later COPAN FLOQSwabs® [COPAN Diagnostics, Murrieta, CA, USA]) was rotated against the tonsil through 360° four times and placed in Thermo Fisher M4RT (Thermo Fisher, Waltham, MA, USA) viral transport medium (3 mL). Swabs were transferred at 4–8°C, aliquoted, and then frozen at –80°C within 48 h.

On day 0, following randomisation, four separate swabs (two swabs from each tonsil) were taken. Separate swabs from each tonsil were then taken once daily from day 1 to day 7, on day 10, and on day 14 (total of 22 swabs). Each swab was processed and tested separately. Vital signs were recorded three times daily by the patient (initial vital signs on the first day were recorded by the study team), and symptoms and any adverse effects were recorded daily. The TaqCheck SARS-CoV-2 Fast PCR Assay (Applied Biosystems, Thermo Fisher Scientific, Waltham, MA, USA) quantitated viral density (RNA copies per mL). This multiplexed real-time PCR method detects the SARS-CoV-2 N and S genes, and human RNase P gene in a single reaction. RNase P provides a measure of the human cell content in the swab eluate, thus allowing for adjustment for variation in intra-cellular viral RNA. Whole-genome sequencing was performed to identify viral variants and allocate genotypes (appendix pages 2-4).

#### Drugs evaluated

The drugs or monoclonal antibodies evaluated in the platform were ivermectin (until 11^th^ April 2022); remdesivir (until 10^th^ June 2022); casirivimab/imdevimab (Thailand only, until 20^th^ October 2022); favipiravir (until 30^th^ October 2022); molnupiravir (until 22^nd^ February 2023); fluoxetine (until 8^th^ May 2023, data not included in this analysis); tixagevimab/cilgavimab (until 4^th^ July 2023, data not included in this manuscript); nitazoxanide (Brazil, Laos and Pakistan, from 18^th^ January 2022, ongoing); ensitrelvir (Thailand and Laos only, from 17^th^ March 2023, ongoing); and ritonavir-boosted nirmatrelvir (from 6^th^ June 2022, ongoing as positive control). All treatment doses were either directly observed or video observed.

### Statistical analysis

#### Data pre-processing

Oropharyngeal eluate viral densities were quantified by PCR on 96-well plates. Each plate contained 10 or 12 ATCC controls (Manassas, VA, USA; these are heat-inactivated SARS-CoV-2 viruses [VR-1986HK strain 2019-nCoV/USAWA1/2020]) varying from 10 to 10^6^ copies per mL. We fitted a linear mixed-effects model to all ATCC control data from all available plates (using R package *lme4* version 1.1.34 [21]) with the genome copies per mL on the log_10_ scale (i.e., a linear relationship between CT values and known log_10_ genomes per ml). The model included fixed effects on the slope and intercept by laboratory (reference laboratory was Thailand), and random effects on the slope and intercept by plate. Visual checks were done to make sure that controls on all plates were in a reasonable range. The mixed-effects model was then used to transform the observed CT values for the oropharyngeal eluates into log_10_ genomes per mL. A CT value of 40 was considered left censored and the plate specific censoring value was used in subsequent analyses. Appendix page 7 shows the estimated standard curves and the model residuals by laboratory.

#### Baseline viral densities

The baseline viral density was defined as the geometric mean of the oropharyngeal eluate SARS CoV-2 densities from the four independent swabs taken before randomisation. Temporal trends in the baseline viral densities were characterised using generalised additive models (GAM) with penalised splines, as implemented in the *mgcv* package version 1.9.0 [22]. As the timing of patient recruitment relative to their onset of symptoms could have also changed over time (and this could affect baseline viral densities), the temporal effect was stratified by the reported number of days since symptom onset. Pearson correlation coefficients between baseline covariates were estimated using the R function *cor.test*.

#### Bayesian hierarchical model to estimate viral clearance rates

The analysis of the serial viral density data used the same core analytical model as in previous publications [5, 13–16]. We characterised oropharyngeal viral clearance under a single exponential decay model (linear decay on the log scale). Under this model, the rate of viral clearance is defined as the slope parameter of a linear fit to the serial log viral density measurements.

The general model likelihood takes the following form:

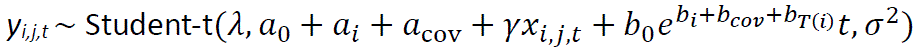

where:

- *y*_i,j,t_ is the log viral density (log_10_ genomes per mL) for the *j*-th swab of patient *i* at time *t*.
- *T*(*i*) is the randomized treatment allocation for individual *i*.
- *σ*^2^ is the variance of the residual error in the model fit; *λ* is the degrees of freedom for the Student-t error model.
- *a*_0_ and *b*_0_ are the population mean intercept (baseline viral density) and slope (viral clearance rate), respectively.
- *a*_i_ and *b*_i_ are the individual random effects on the intercept and slope, respectively.
- *a*_cov_ and *b*_cov_ are the linear covariate effects on the intercept and slope, respectively.
- *x*_i,j,t_ is the human RNase P CT (scaled to have mean 0) for the *j*-th swab from patient *i* at time *t*, with a *γ* parameter adjusting for the effect of human RNase P on the estimated viral density in the oropharyngeal eluates.

Covariate terms for the slope and intercept were the reported days since symptom onset, study site, age, sex, and number of vaccine doses received. This model parameterised the treatment effect relative to a reference intervention (e.g. no study drug) as a proportional change (*e^bT^*^(*i*)^). As a sensitivity analysis, we parameterised the treatment effect as an additive change (*b*_0_*e*^*bi*+*bco*ʋ^ + *b*_*T*(*i*)_). Model comparison was done using leave-one-out as implemented in the package *loo* version 2.6.0 [23].

All viral densities below the lower limit of quantification (defined as a CT value of 40) were treated as left-censored (the likelihood is the integral of the likelihood function below the censoring value). The linear model fitted to data between days 0 and *T*_max_ (*T*_max_ was 7 days in the primary analysis) estimates the average clearance rate over *T*_max_ days. We denote this as *α*_0−Tmax_.

All models were fitted using weakly informative priors on all parameters (appendix pages 5-6). These priors help computational convergence but have no effect on the parameter estimates (we showed this in previous analyses [13]). In previous analyses we also used a nonlinear ‘up-down’ model (linear increase followed by linear decrease), but this also had no effect on treatment effect estimates [13].

#### Temporal changes in viral clearance dynamics

To assess the temporal changes in viral clearance we added a penalised B-spline of degree 4 to the population mean intercept *a*_0_ (baseline viral density) and population mean slope *b*_0_ (population viral clearance rate) in the reference arm (for most analyses this is the no study drug arm). This was done by having many knots at regular intervals (20 knots in the main analysis) with an informative penalisation prior on parameter changes across knots. The penalisation prior governs the smoothness of the spline fit (https://github.com/milkha/Splines_in_Stan).

#### Meta-analysis of treatment effects

The interventions studied (ivermectin, remdesivir, favipiravir, molnupiravir, ritonavir-boosted nirmatrelvir, and casirivimab/imdevimab) were not randomised concurrently (appendix page 8). Thus, the large observed temporal changes in baseline viral clearance rates bias cross-comparisons. We adjusted for temporal confounding by explicitly incorporating into the model the temporal changes in viral clearance rates in the treatment effect estimation. We fitted the full Bayesian linear model with a spline term on the baseline clearance rate in the no study drug arm (which spans the entire study period) as a function of the calendar date, with treatment effects parameterised as proportional changes in the average clearance. As a sensitivity analysis, we assessed treatment effect heterogeneity with respect to the SARS-CoV-2 major lineages. This was done by incorporating interaction terms between the intervention and the and viral lineage.

#### Optimal design

In light of the substantial changes observed in the viral clearance rates *α*_0−7_ in COVID-19 over the past two years, we used the available comparative data to assess the optimal trial design for pharmacometric assessment and thus the rapid identification and evaluation of effective antivirals. We define ‘optimal’ as the design (duration and frequency of sampling) which maximises the expected z-score for differences in viral clearance rates when comparing an effective randomised intervention with the concurrent no treatment arm or comparing two concurrently randomised interventions with different antiviral efficacies. The z-score is the estimated effect size divided by the estimated standard error. We bootstrapped the data (sampling patients in each comparison with replacement) to obtain uncertainty intervals for the z-score estimates for each comparison. In order for the z-scores to be comparable, each bootstrap sample contained 50 patients per arm.

The following designs were compared:

- Varying the duration of follow-up from 2 days (i.e., using qPCR measurements taken on days 0, 1 and 2) to 14 days (using all available qPCR data);
- Varying the number of swabs taken each day (1 or 2);
- Comparing twice daily swabs taken on days 0 to 4 (10 qPCR measurements), versus twice daily every other day (0, 2, and 4: 6 qPCR measurements), versus twice daily only on days 0 and 4 (4 qPCR measurements).

Empirical expected z-scores were estimated under the linear model for six separate intervention comparisons (each comparison used concurrently randomised patients): remdesivir versus no study drug; casirivimab/imdevimab versus no study drug; molnupiravir versus no study drug; ritonavir boosted nirmatrelvir versus no study drug; and ritonavir-boosted nirmatrelvir versus molnupiravir. The data for ritonavir-boosted nirmatrelvir versus no study drug spanned 16 months with a brief hiatus in recruitment from January to February 2023. We therefore arbitrarily split these data into two separate comparisons, before January 2023 and after February 2023. This allowed assessment of how much the temporal change in viral clearance was driving the observed results. For each of these six comparisons, and each sampling design (duration of follow-up and number of samples), we bootstrapped the data 50 times (sampling the patients with replacement) and fitted the linear model to estimate the treatment effect and standard error.

### Analysis code

All Bayesian models are written in stan and fitted using the *rstan* interface version 2.32.3 [24]. All analyses were done using R programming language version 4.3.2 [25].

## Results

### Patient cohort

Between the 29^th^ September 2021 and 20^th^ October 2023, 1262 patients were randomised in the PLATCOV trial across six sites in four countries (Thailand, Brazil, Pakistan, and Laos). Patients randomised to ensitrelvir, the combination treatment of ritonavir-boosted nirmatrelvir and molnupiravir, and nitazoxanide were not included in this analysis as these comparisons are ongoing and their data are still blinded (Figure 1). Patients randomised to fluoxetine and tixagevimab/cilgavimab were not included as their data had not been published at the time of the analysis [26]. After excluding patients who withdrew consent, or who were not SARS-CoV-2 positive on any follow-up samples, or who had fewer than 2 days follow-up, the analysis population consisted of 800 patients randomised across seven arms (not all concurrently, see appendix page 8). Nearly all included patients had received at least one vaccine dose (>98%) and the majority were fully vaccinated (>88%) prior to symptom onset (Table 1; appendix page 9). The majority (85%) of patients were randomised at one site in Thailand (Hospital for Tropical Diseases, Bangkok). The mean time from symptom onset to randomisation was 2.1 days (standard deviation [SD]: 0.8) and the geometric mean baseline viral density in oropharyngeal eluates was approximately 5.5 log_10_ genomes per mL (SD: 1.2). Nearly all patients had complete viral density data between days 0 and 7 (less than 5% missing data in all intervention arms, see appendix page 10).

**Figure 1:**
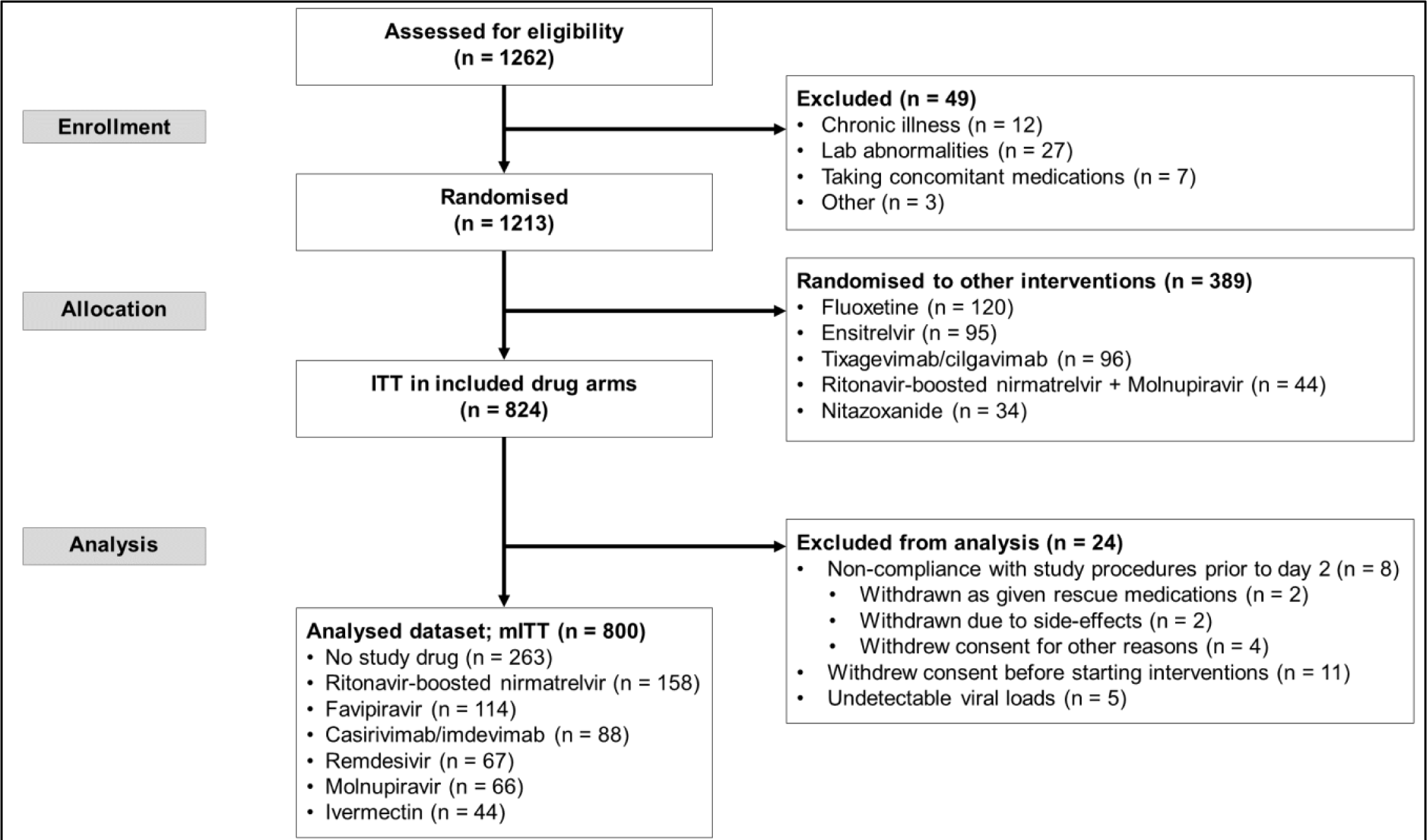
PLATCOV trial profile and selection of patients used in this analysis. This analysis includes patients enrolled between 30th September 2021 and 20th October 2023 who met the modified intention to treat (mITT) criteria and whose viral clearance data have been unblinded and published. Patients are only excluded from the mITT if protocol deviations occur on days 0 to 2.

**Table 1:** Baseline demographics for the 800 patients included in the analysis. Unless specified, data are shown as *n* (%); ^†^mean (standard deviation); ^‡^median (interquartile range).

|  | No study drug<br>(n=263) | Ritonavir-boosted<br>nirmatrelvir<br>(n=158) | Favipiravir<br>(n=114) | Casirivimab/<br>imdevimab<br>(n=88) | Remdesivir<br>(n=67) | Molnupiravir<br>(n=66) | Ivermectin<br>(n=44) |
| --- | --- | --- | --- | --- | --- | --- | --- |
| Brazil | 26 (10) | 4 (3) | 16 (14) | 0 | 9 (13) | 0 | 0 |
| Thailand | 230 (88) | 150 (95) | 98 (86) | 88 (100) | 58 (87) | 65 (99) | 44 (100) |
| Laos | 3 (1) | 4 (3) | 0 | 0 | 0 | 1 (2) | 0 |
| Pakistan | 4 (2) | 0 | 0 | 0 | 0 | 0 | 0 |
| Age (years)† | 31.1 (8.1) | 31.3 (8.9) | 30.2 (7.5) | 27.9 (7.3) | 30.1 (8.2) | 31.3 (7.5) | 30.0 (7.0) |
| Female | 173 (66) | 109 (69) | 71 (62) | 55 (62) | 35 (52) | 37 (56) | 24 (55) |
| Weight<br>(kg)† | 63.0 (13.6) | 61.5 (12.3) | 63.0 (13.6) | 60.4 (12.3) | 63.9 (11.0) | 63.4 (14.7) | 61.6 (12.3) |
| BMI† | 23.2 (4.1) | 23.0 (3.8) | 23.1 (3.7) | 22.1 (3.1) | 22.7 (3.1) | 23.1 (4.0) | 22.3 (3.2) |
| Interval since<br>symptom<br>onset (days)‡ | 2 (1-3) | 2 (1-2) | 2 (2-3) | 2 (2-3) | 2.5 (2-3) | 2 (2-2) | 2 (2-3) |
| Admission viral<br>load (log<sub>10</sub><br>genomes/mL) ‡ | 5.6 (4.7-6.4) | 5.6 (4.6-6.4) | 5.5 (4.7-6.2) | 5.7 (5.0-6.4) | 5.3 (4.8-6.3) | 5.8 (5.0-6.4) | 5.6 (5.0-6.6) |
| Vaccinated (%) | 253 (96) | 153 (97) | 112 (98) | 85 (97) | 64 (96) | 65 (99) | 43 (98) |
| Start date | Sep 29, 2021 | Jun 5, 2022 | Oct 10, 2021 | Oct 1, 2021 | Oct 4, 2021 | Jun 5, 2022 | Sep 29, 2021 |
| Finish date | Oct 20, 2023<br>(ongoing) | Oct 20, 2023<br>(ongoing) | Oct 29, 2022 | Oct 19, 2022 | Jun 7, 2022 | Feb 14, 2022 | Apr 11, 2022 |
| SARS-CoV-2<br>variants |  |  |  |  |  |  |  |
| Delta | 10 (4) | 0 | 11 (10) | 13 (15) | 10 (15) | 0 | 12 (27) |
| BA.1 | 13 (5) | 0 | 21 (18) | 15 (17) | 20 (30) | 0 | 14 (32) |
| BA.2 | 52 (20) | 1 (1) | 42 (37) | 30 (34) | 37 (55) | 5 (8) | 18 (41) |
| BA.2.75 | 43 (16) | 30 (19) | 5 (5) | 5 (6) | 0 | 28 (42) | 0 |
| BA.4 | 2 (1) | 3 (2) | 3 (3) | 0 | 0 | 2 (3) | 0 |
| BA.5 | 42 (16) | 26 (17) | 32 (28) | 25 (28) | 0 | 28 (42) | 0 |
| XBB | 29 (11) | 22 (14) | 0 | 0 | 0 | 3 (5) | 0 |
| XBB.1.5-like | 67 (26) | 75 (48) | 0 | 0 | 0 | 0 | 0 |
| Other | 5 (2) | 1 (2) | 0 | 0 | 0 | 0 | 0 |

### Baseline viral densities

The baseline oropharyngeal eluate viral densities remained high over the two-year period (appendix page 11) but there were systematic trends over time associated with different SARS-CoV-2 variants. The reported interval since symptom onset was negatively correlated with the baseline viral density (correlation coefficient *ρ* = –0.22 [95% confidence interval (CI): –0.29 to –0.16]; *R*^2^ = 0.05) (Figure 2A). Each reported additional day since symptom onset corresponded to a 1.9-fold [95% Crl: 1.5 to 2.4] decrease in the baseline viral density; and males had 1.4-fold [95% Crl: 1.0 to 2.1] higher baseline viral load densities (appendix page 12). There were small changes in the mean reported number of days since symptom onset over time. For example, during the Omicron BA.1 wave (1^st^ January 2022 to 11^th^ March 2022) patients were recruited slightly later on average, Figure 2B. In a multivariable spline model stratified by the interval since symptom onset, there was evidence of systematic temporal changes in baseline viral density over time which were not explained by differences in time from symptom onset (Figure 2C). As these are observational data it is not possible to determine causality (e.g., whether these differences result from variant specific mutations in the spike protein) but the data are compatible with higher peak viral densities with specific variants such as BA.2 and XBB.1.5-like.

**Figure 2:**
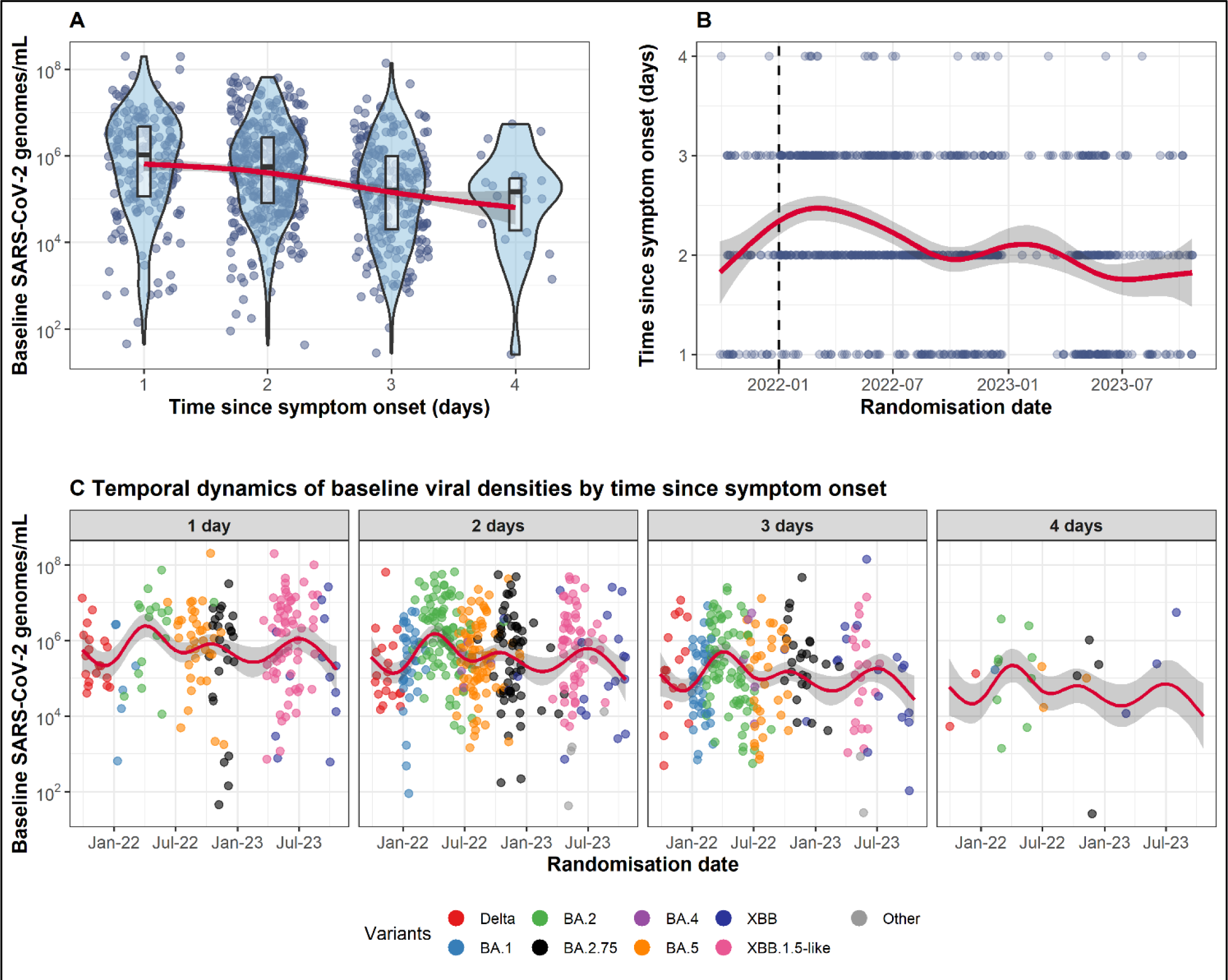
Changes in symptoms duration at enrollment and baseline viral densities over a two year period (2021-2023). Panel A: relationship between reported time since symptom onset and baseline viral density; panel B: temporal changes in the reported time since symptom onset. The vertical dashed line indicates the first Omicron BA.1 infection enrolled in the study; panel C: temporal changes in the baseline viral density stratified by reported time since symptom onset. Red lines (shaded areas) represent mean estimated values (95% confidence intervals) under a generalised additive model.

### Viral clearance rates over time

Viral clearance increased substantially over the two years of the trial, as shown clearly in patients randomised to no study drug (appendix page 13). Figure 3 shows the individual clearance rate estimates *α*_0−7_ under the hierarchical Bayesian model with a spline term to capture temporal changes. In the no study drug arm, median viral clearance rates have doubled approximately from –0.43 log_10_ units per day in September 2021 (corresponding to a half-life of around 16.6 hours [interquartile range (IQR): 15.3 to 18.2]), to –0.78 log_10_ units per day in October 2023 (half-life of around 9.2 hours; IQR: 8.0 to 10.6). This change corresponds to a relative shortening in viral clearance half-life of 44% [95% credible interval [CrI]: 19 to 64%] over two years. Similar trends were noted for the treated individuals. For example, the mean viral clearance rate in the ritonavir-boosted nirmatrelvir arm increased from –1.12 log_10_ units per day in June 2022 (half-life of 6.4 hours; IQR: 5.7 to 7.3) to –1.50 log_10_ units per day in October 2023 (half-life of 4.8 hours; IQR: 4.2 to 5.5). This change corresponds to a relative shortening in viral clearance half-life of 26% [95%CrI: –4 to 42%]. The reduction of viral clearance half-life was most apparent early in the study between September 2021 (Delta variant) to in mid-February 2022 (BA.2 variant, appendix page 14). Subsequently, the half-life plateaued at around 12.5 hours during the BA.2, BA.4, BA.5, and BA.2.75 variants, and gradually reduced again after the emergence of XBB and XBB.1.5-like variants in January 2023. There was no clear relationship between individual viral clearance rate estimates and the number of days since symptom onset, sex, age, or the number of vaccine doses received (appendix page 12).

**Figure 3:**
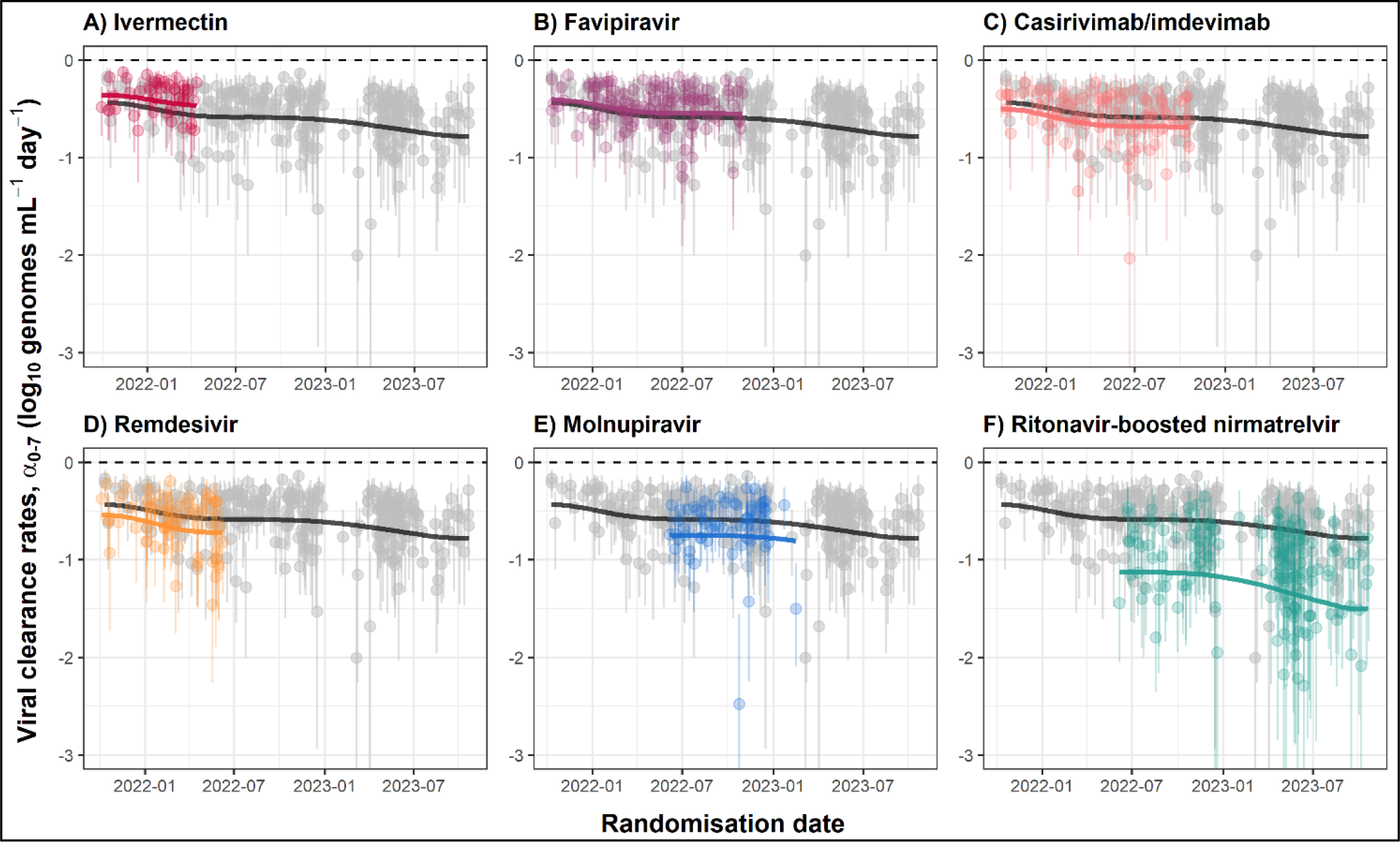
Individual patient data meta-analysis of the platform trial showing estimated rates of viral clearance between days 0 and 7 (α0−7). Average clearance rates for each intervention (coloured lines) and the no study drug arm (black line) are estimated from a spline fit. Treatment effects were parameterised as a proportional change in rate. The grey circles and black lines for the no study drug arm are identical in each panel. Vertical lines show 95% credible intervals under the linear model. A negative sign of the clearance rate indicates a decreasing directional change in viral density.

**Figure 4:**
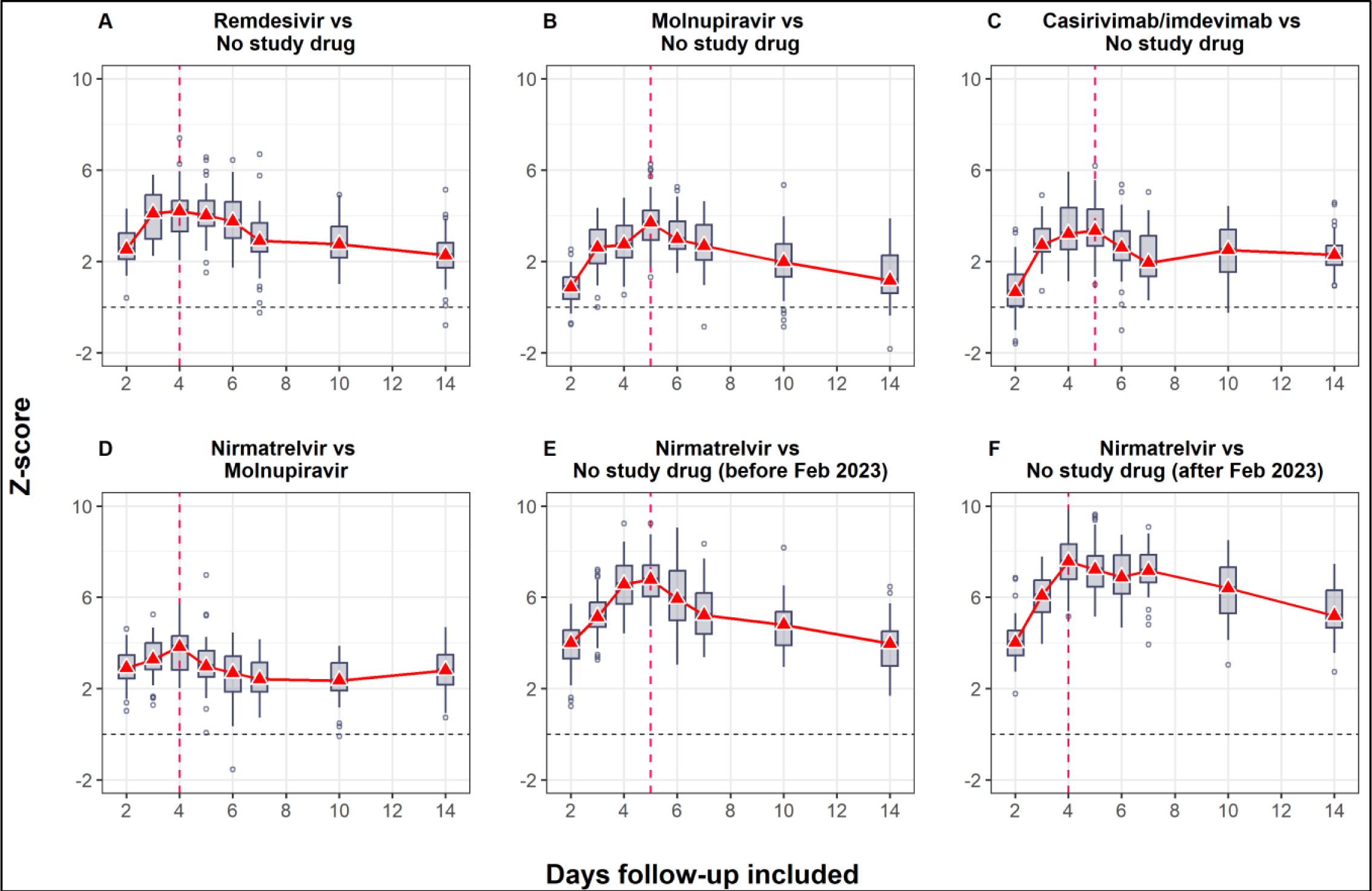
Z-scores for the six treatment effects as a function of the follow-up duration. The boxplots show the median and interquartile range of the z-scores for the 50 bootstrap iterations. Each bootstrap adatset contained 50 patients per arm. The red vertical dashed lines indicate the follow-up durations with maximal z-scores. All comparisons use concurrent controls only.

### Optimising trial design

Figure 4 shows the expected z-scores for 6 randomised comparisons with a sample size of *n*=50 per arm as a function of the duration of follow-up data included in the estimated viral clearance rate (varying from day 2 until day 14). For all pairwise comparisons there was a clear inverted-parabolic relationship between the expected z-score and the duration of follow-up. The expected z-score was maximised for durations between 4 and 5 days. This implies that 4-5 days follow-up is optimal in terms of power when the data are analysed under a linear model framework. Fitting a single component log-linear model over a longer time period systematically reduced the estimate of the slope (i.e., lengthens the half-life) as it incorporated more of the slower *β*-phase (second phase) of viral elimination in the estimate. Additional analyses highlighted the importance of taking duplicate oropharyngeal swabs (appendix page 15). Reducing the frequency of the viral density measurements reduced the expected z-score but with a lesser effect.

### Comparative assessment of antiviral interventions

Under the linear model with adjustment for temporal changes in clearance rates, there was a clear hierarchy between the studied interventions (Figure 5). This hierarchy remained consistent when estimating treatment effects using the average viral clearance rates up until day 5 (*α*_0−5_) or up until day 7 (*α*_0−7_). Ritonavir-boosted nirmatrelvir had the greatest effect on viral clearance rates (approximately 90% increase in average clearance rates *α*_0−7_; approximately 130% increase in average clearance rates *α*_0−5_). The small molecule drugs remdesivir and molnupiravir had very similar effects (approximately 35% increase in average clearance rates *α*_0−7_; approximately 50 to 60% increase in average clearance rates *α*_0−5_). The average treatment effect for the monoclonal antibody casirivimab/imdevimab was of similar magnitude (ignoring known treatment effect heterogeneity [14]). This meta-analysis confirmed the absence of any measurable effect of high-dose ivermectin or high-dose favipiravir. For all four effective interventions, the analysis using the *α*_0−5_ average clearance rates estimated substantially larger effect sizes, albeit with slightly wider uncertainty intervals. There was no evidence of treatment effect heterogeneity for the small molecule drugs by viral variants, whereas the effect of casirivimab/imdevimab varied considerably across the major viral variants (appendix page 16).

**Figure 5:**
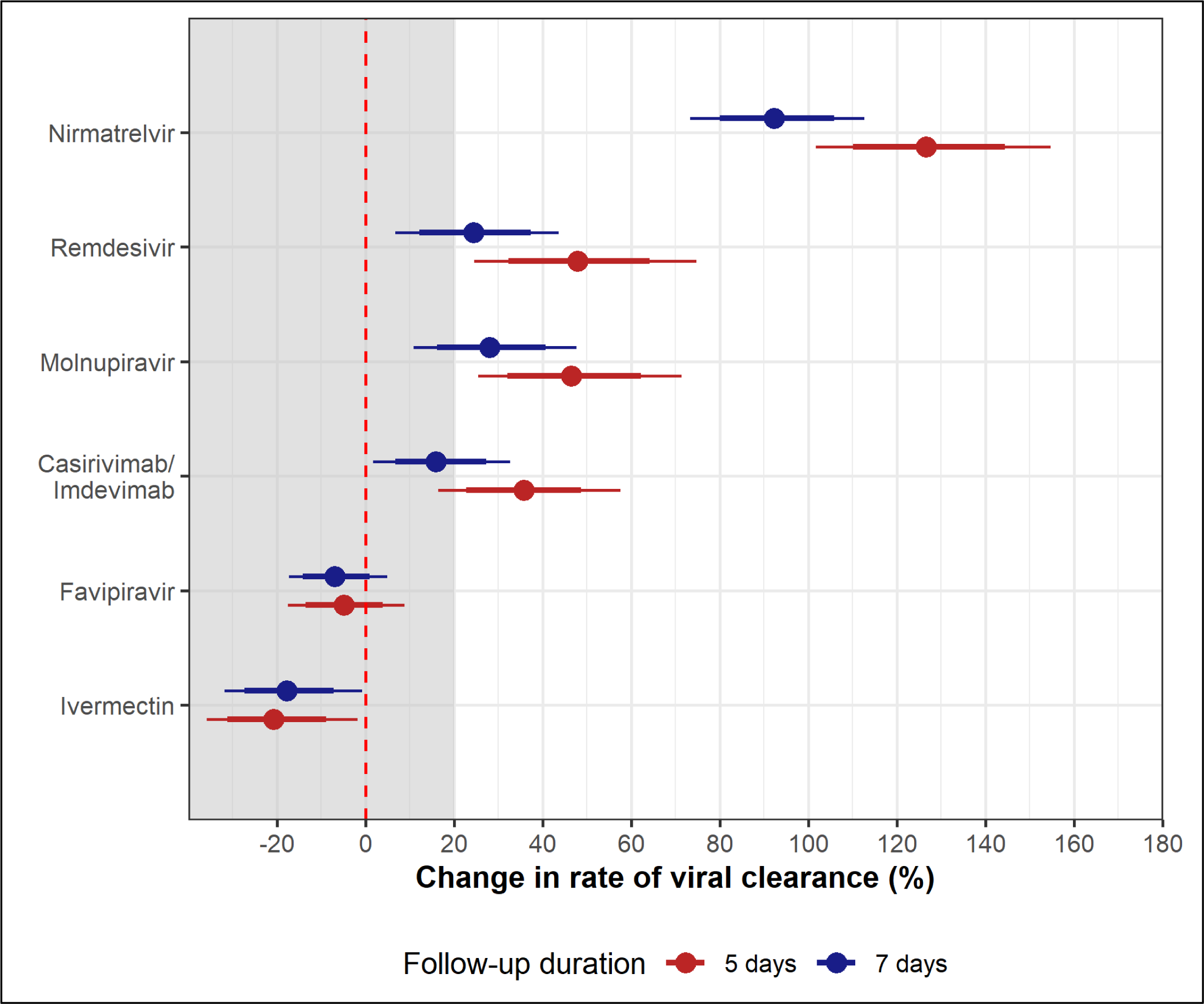
Individual patient 6data meta-analysis of the treatment effect of the six randomised interventions relative to no study drug. Red: treatment effects based on the average clearance rates over 5 days (*α*_0−5_); blue: treatment effects based on the average clearance rates over 7 days (*α*_0−7_). The models explicitly adjusted for temporal changes in viral clearance in the no study drug arm using penalised B-splines. Points: median posterior estimate; thick and thin lines: 80% and 95% credible intervals, respectively.

## Discussion

SARS-CoV-2 oropharyngeal clearance rates in uncomplicated COVID-19 infections have become substantially faster over the past two years. Natural viral clearance is now twice as fast as it was in September 2021. This very granular prospectively gathered dataset confirms the findings of other larger scale observational cohorts [27]. In this studied cohort, most of whom were fully vaccinated, waves of different viral variants succeeded each other, following a generally similar pattern to that observed in most areas of the world. There was no clear association between particular viral variants and increases in viral clearance rates. Instead, there appears to have been a steady increase in clearance rates across all variants over time. Some variants (e.g., BA.2.75) were clearly associated with higher baseline viral densities, which was not explained by differences in the interval from symptom onset. It is not possible to ascribe with confidence the underlying cause for these higher baseline viral loads, but it would be compatible with either differences in viral replication [28] or differences in tropism [29].

The substantial acceleration in natural viral clearance over the past two years presumably reflects the interplay between the acquisition of immunity and the antigenic changes in the evolving variants. This acceleration in natural viral clearance has important implications for the assessment of in-vivo antiviral activity. SARS-CoV-2 oro/nasopharyngeal clearance is biphasic [17–19]. Effective drugs substantially accelerate the first phase. Two years ago, when viral clearance rates were much slower, the inflexion in the clearance curve (transition from the first to the second slower phase) was close to seven days, so fitting a single rate constant to the log-linear decline in viral densities over seven days incurred relatively little bias. At current rates of viral clearance, the inflexion point is much earlier, so forcing a single rate constant to the serial qPCR values over seven days incurs greater bias resulting in progressive underestimation of the initial phase rate of clearance. This is important for historical comparisons of antiviral activity as, with any viral clearance measure, drugs today will result in faster viral clearance than they did earlier in the pandemic. Moderately effective drugs evaluated two years ago (e.g., remdesivir) resulted in viral clearance rates that are similar to those in the no treatment arm of the study today.

The PLATCOV study has characterised the effect of several antiviral drugs with findings which are generally consistent with earlier clinical trials assessing their efficacy in the prevention of disease progression. Comparative estimates of in vivo antiviral activity allow for rational selection of drugs now that comparison based on clinical endpoints is no longer possible because of the prohibitively large sample sizes required in clinical trials. Using the observed differences in the viral clearance profiles between effective and ineffective drugs or the no treatment arm allowed determination of the sampling duration which best characterised these differences. The greatest differences between effective and ineffective (or no) drugs were observed for assessments made from serial samples taken over 4-5 days. Although there is substantial inter-individual variation in clearance rates, and also intra-individual variation between the serial viral density estimates, with current viral clearance rates daily sampling still has adequate discriminatory power. But, if this trend of increasing rapidity of viral clearance continues, then it may be necessary to sample twice daily over a shorter period. Shortening the viral clearance serial sampling to five days simplifies the comparative assessment of antiviral drugs in COVID-19 (although later sampling is still necessary if rebound is being assessed).

These data emphasise the critical importance of fixed ratio contemporary comparators in platform trials. Temporal confounding across non-currently randomised interventions or for time varying randomisation ratios (this occurs in response adaptive trials) requires model dependent adjustment. Even an ineffective drug will appear effective if compared with a historical control. The exact ranking of all unblinded interventions studied on the platform in the meta-analysis is dependent in part on correct adjustment for the temporal trends. This issue is particularly salient for the comparison between remdesivir and molnupiravir.

SARS-CoV-2 is today predominantly a mild infection in vaccinated individuals which does not require specific antiviral treatment. This justifies the recruitment of patients into the pharmacometric assessment who receive no specific treatment. But in patients with underlying conditions or the elderly, COVID-19 is still potentially dangerous and specific antiviral treatment is required. There is no reason to believe that antiviral activities are different in these high-risk subgroups to those observed in low-risk patients enrolled in this study. At the beginning of the pandemic there were no effective interventions and so identifying minor accelerations in viral clearance was relevant. Today modest acceleration in the rate of viral clearance may still be therapeutically relevant for chemoprevention [26], but it is very unlikely that less effective drugs than those now being used would be deployed for the treatment of symptomatic COVID-19. The simple methodology employed in the PLATCOV trial is efficient, and very well tolerated, and it identifies efficacious antivirals (i.e., those which result in viral clearance rates that are >20% faster than no drug) with sample sizes which are usually less than 40 studied patients per arm.

Although this is the largest detailed pharmacometric study in COVID-19, it has some limitations. Most of the patients were studied in Bangkok, Thailand so the temporal trends observed could be different in other parts of the world. The cause of the substantial inter-patient variations in viral clearance and the overall acceleration in viral clearance over the past two years has not been characterised adequately. Over 95% of patients were vaccinated before the enrolment infection and so we cannot characterise differences in treatment effects between vaccinated and unvaccinated individuals. Although there is a clear rationale for using viral clearance as a surrogate endpoint in assessing therapeutics [8, 9], additional data are still needed to characterise the relationship between acceleration in viral clearance and clinical outcomes such as rate of symptom clearance.

In summary, SARS-CoV-2 viral clearance has accelerated substantially over the past two years necessitating a shortening of the sampling time to evaluate and compare antiviral drugs efficiently.

## Data Availability

All data and code necessary to reproduce the results in this analysis are openly available at https://github.com/jwatowatson/Determinants-viral-clearance

## Acknowledgements

NJW is a Principal Research Fellow funded by the Wellcome Trust (093956/Z/10/C). JAW is a Sir Henry Dale Fellow funded by the Wellcome Trust (223253/Z/21/Z). This study is supported by Wellcome Trust grant reference 223195/Z/21/Z through the COVID-19 Therapeutics Accelerator. We thank all the patients with COVID-19 who volunteered to be part of the study. We thank the data safety and monitoring board (Tim Peto, André Siqueira, and Panisadee Avirutnan); the trial steering committee (Nathalie Strub-Wourgaft, Martin Llewelyn, Deborah Waller, and Attavit Asavisanu); Sompob Saralamba and Tanaphum Wichaita for developing the Rshiny randomisation app; and Mavuto Mukaka for invaluable statistical support. We also thank all the staff of the clinical trials unit at MORU, Thermo Fisher for their excellent support with this project, and all the hospital staff at the Hospital for Tropical Diseases, as well as those involved in sample processing in MORU and the processing and analysis at the Faculty of Tropical Medicine, Mahidol University, molecular genetics laboratory, and the malaria laboratory. We thank the Department of Medical Services, Ministry of Public Health, Thailand for the generous support and the donation of the molnupiravir and ritonavir-boosted nirmatrelvir for this study. We thank the MORU Clinical Trials Support Group for data management, monitoring and logistics, and the purchasing, administration and support staff at MORU.

This research was partly funded by Wellcome. A CC BY or equivalent licence is applied to the author accepted manuscript arising from this submission, in accordance with the grant’s open access conditions.

## Authorship contributions

PW: conceptualisation, data curation, formal analysis, methodology, visualisation and writing original draft; WHKS: conceptualisation, funding acquisition, investigation, methodology, project administration, supervision, validation and writing-original draft; NJW: conceptualisation, funding acquisition, methodology, supervision, validation and writing-original draft; JAW: conceptualisation, data curation, formal analysis, funding acquisition, methodology, visualisation and writing original draft; PJ: investigation, methodology, project administration, supervision, and validation; SB, and SS: investigation, methodology and project administration; VL, TS, TN, BH, and KP: investigation, methodology, and supervision; EMB: data curation, and formal analysis; JK, WPa, and PK: investigation, methodology; MM, and EAA: investigation, supervision, funding acquisition; KC and MI: formal analysis, investigation, resources, supervision; SP, AMD, MMT, WPi, WPh: methodology, investigation, resources, supervision; NPJD: funding acquisition, methodology, investigation, resources, supervision. All authors were involved in writing, review, and editing of the manuscript. PW, WHKS, EMB, MI, NJW, JAW have directly accessed and verified the underlying data reported in the manuscript. All authors had full access to all the data in the study and had final responsibility for the decision to submit for publication.

## Data Sharing

All code and de-identified participant data required for replication of the study’s endpoints are openly accessible via Zenodo, as well as the study protocol and statistical analysis plan, from publication date onwards. Individual patient data can be requested and may be shared according to the terms defined in the MORU data sharing policy with other researchers to use in the future from the date of publication. Further information on how to apply is on the MORU Tropical Health Network site.

